# Antimicrobial stewardship strategies to optimise antibiotic prescribing in primary care: physicians’ awareness, beliefs, and strategy use

**DOI:** 10.64898/2026.11.12.26367755

**Authors:** Aleksandra J. Borek, Iga Palacz-Poborczyk, Julia Kuzminska, Aleksandra Luszczynska, Karolina Kłoda, Mateusz Babicki

## Abstract

**Background:** National efforts to promote antimicrobial stewardship (AMS) and prevent antimicrobial resistance (AMR) in primary care need strengthening. Research supports clinician-targeted AMS strategies to optimise antibiotic prescribing, but implementing them in clinical settings is challenging.

**Aim:** Assessment of primary care physicians’ awareness of, use of, and beliefs about evidence-based AMS strategies, and perceived barriers and facilitators to implementing them in a country with high antibiotic consumption rates (Poland).

**Methods:** A pre-registered observational survey study. Eligible physicians worked in primary care in Poland for 6+ months and were recruited via professional networks and social media. Data were collected through a pre-piloted online questionnaire and analysed descriptively.

**Results:** 270 physicians (68.2% Specialists/Residents in Family Medicine) participated. Respondents were aware of the threat of AMR (88%), the importance of AMS (93%) and their role in AMS (89%). They most recognised, utilised, perceived as effective, and recommended rapid diagnostic tests, clinical guidelines, and scales/criteria. Respondents reported high familiarity but lower engagement with communication strategies, national actions, education/training, and campaigns. Delayed prescriptions were the least used and recommended. Main barriers were time constraints and insufficient awareness of which strategies to implement. Recommended strategies included tests, guidelines, scales/criteria, AMS action plans for general practices, patient communication resources, and promoting awareness of AMS and its role in high-quality care.

**Conclusion:** Awareness and use of evidence-based AMS strategies among physicians working in Polish primary care clinics appear high. Improvements may require reaching disengaged physicians, increasing uptake of underutilised strategies, and strengthening the impact of already used strategies.

## Introduction

Optimal antibiotic prescribing helps prevent antimicrobial resistance (AMR) and adverse patient outcomes. This is particularly important in primary care, where most antibiotics are used [1], often for self-limiting acute respiratory tract infections (RTIs). Antimicrobial stewardship (AMS) strategies aim to promote optimal antibiotic use, that is, only when necessary, with the right drug, at the right time and dose, and for the right duration [2]. However, progress towards AMS-related targets in Europe has been poor, prompting repeated calls to strengthen national AMS programmes [1].

Several types of clinician-targeted AMS strategies aim to optimise antibiotic prescribing in primary care: educational materials, meetings and outreach visits; audit and feedback; reminders (including prompts and decision support systems); financial interventions; point-of-care tests (POCTs); communication strategies (for clinician-delivered patient education, improved clinician-patient interaction, and shared decision-making); clinician-targeted mass media campaigns; and delayed prescriptions [3]. Evidence supports the benefits of POCTs [3–5], communication skills training [3,5], educational interventions [6,7], audit and feedback [8], clinical decision support systems [9], financial interventions [10], and delayed prescriptions [11] in reducing antibiotic prescribing/use in primary care. However, while these strategies work in research settings, their implementation, sustainable use and impact in routine care are often neglected [12,13]. These outcomes are affected by key psychological determinants of behaviour: capability (e.g., knowledge), motivation (e.g., beliefs about capabilities and consequences), and opportunity (e.g., environmental context) [14]. Understanding these determinants helps identify ways to improve the use of evidence-based AMS strategies.

Poland is a high-income country with antibiotic consumption rates above the European Union (EU) average and the EU targets, and with high use of broad-spectrum antibiotics in the community [1,15]. Meeting the EU targets would require a 27% reduction in overall antibiotic consumption between 2019 and 2030, compared with the 4% change observed until 2024 [1]. The National Health Programme 2021–2025 includes a task to prevent AMR, but without specifying how to achieve this in primary care or the corresponding outcome measure [16]. Thus, effort is needed to identify the factors driving antibiotic over-prescribing and ways to address this challenge in Polish primary care.

Previous surveys of physicians in Poland (not specific to primary care) found high levels of knowledge about antibiotics and AMR [17,18], but low awareness of information resources and national campaigns related to antibiotics/AMR [19]. A survey of primary care physicians showed that physicians perceived their decisions about whether to prescribe antibiotics as most influenced by clinical factors: physical examination, recommendations, and tests [20]. However, it remains unclear which evidence-based AMS strategies have been used in the Polish primary care context, and how to address not only capability (knowledge and skills) but also motivational and contextual factors [14], as well as barriers and facilitators to using AMS strategies [12]. For example, studies in England identified widespread implementation and a positive impact of monitoring, auditing, and feedback on antibiotic prescribing, together with prescribing targets [21,22].

The study aimed to examine physicians’ awareness of, use of, and beliefs about evidence-based AMS strategies, as well as perceived barriers, facilitators and priorities related to their use. It forms part of a larger project to identify opportunities to improve antibiotic prescribing and strengthen antimicrobial stewardship in Polish primary care.

## Methods

This was an observational, cross-sectional, online survey study. It was prospectively registered (https://doi.org/10.17605/OSF.IO/V8HRT) and reported following a relevant checklist (Supplement 1)[23].

A questionnaire was designed by listing clinician-targeted AMS strategies in primary care, drawing on the Cochrane systematic review [3]. Next, survey questions related to the AMS strategies that are known to be used/available in Polish primary care were drafted and refined by the study team (including general practitioners) to ensure relevance, accuracy, and clarity. The questionnaire was transferred online and piloted by the study team (two general practitioners, three social scientists) and by four external physicians meeting our respondent eligibility criteria (1 general practitioner, 2 paediatricians, 1 physician without a specialisation). We conducted three rounds of piloting and refinements until we received no further suggestions. The questionnaire took 10-15 minutes to complete.

The final questionnaire (Supplement 2) comprised 23 questions (plus sub-questions/items) across five sections. Following (1) screening and consent and (2) respondent characteristics (Table 1), section (3) covered three items on the perceived role in AMS, the importance of AMS, and the risk of AMR in primary care (5-point scale from strongly agree to strongly disagree; Figure 1). Section (4) addressed seven types of AMS strategies: (i) clinical guidelines and recommendations, (ii) information/educational campaigns, (iii) education and training, (iv) clinical scales/criteria and diagnostic algorithms, (v) rapid diagnostic tests (e.g., C-Reactive Protein [CRP], strep tests, combo antigen tests), (vi) specific communication strategies about antibiotics (e.g., discussing patient expectations, antibiotic side effects, typical symptom duration), and (vii) delayed antibiotic prescriptions. Respondents were asked four items for each strategy, covering familiarity (awareness), self-reported use (behaviour), confidence in use (motivation/beliefs about capabilities), and perceived effectiveness (motivation/beliefs about consequences), with four response options (agree, partly agree, disagree, not relevant; Figure 2). Section (5) addressed the implementation of AMS strategies. Respondents were asked multiple-choice questions about which of the available AMS strategies should be promoted and used more, and which of the potential AMS strategies (not currently used) could positively impact antibiotic prescribing (Figure 3). They were also asked multiple-choice questions about the perceived main barriers and facilitators to implementing AMS (Figure 4). Space for optional free-text comments was provided in each section. Except for the required sections 1 and 2, all other sections and questions were optional to increase respondent retention.

**Figure 1.**
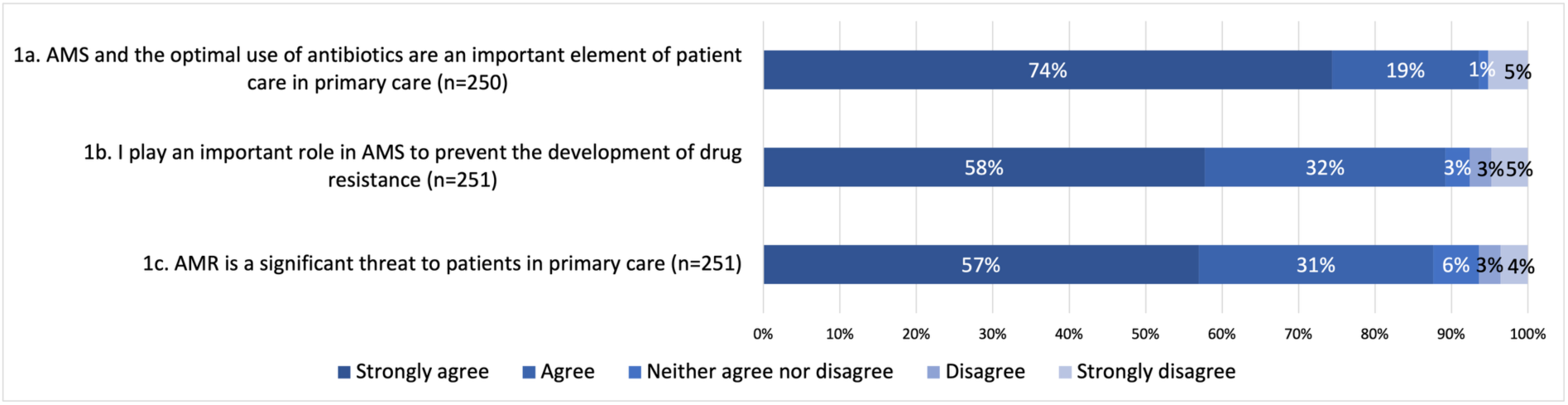
Perceived importance of and role in AMS.

**Figure 2.**
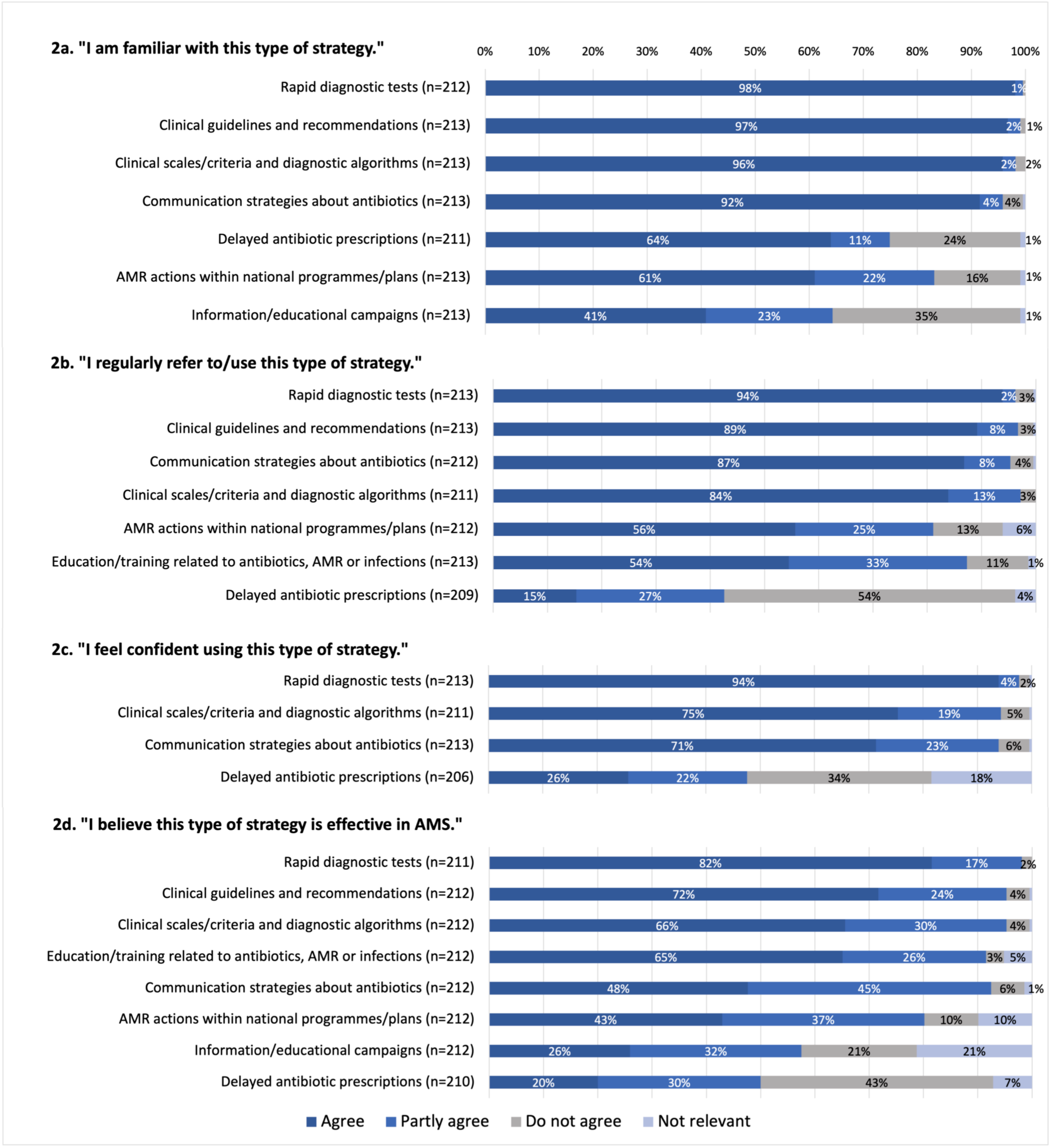
Experiences of and views on AMS strategies.

**Figure 3.**
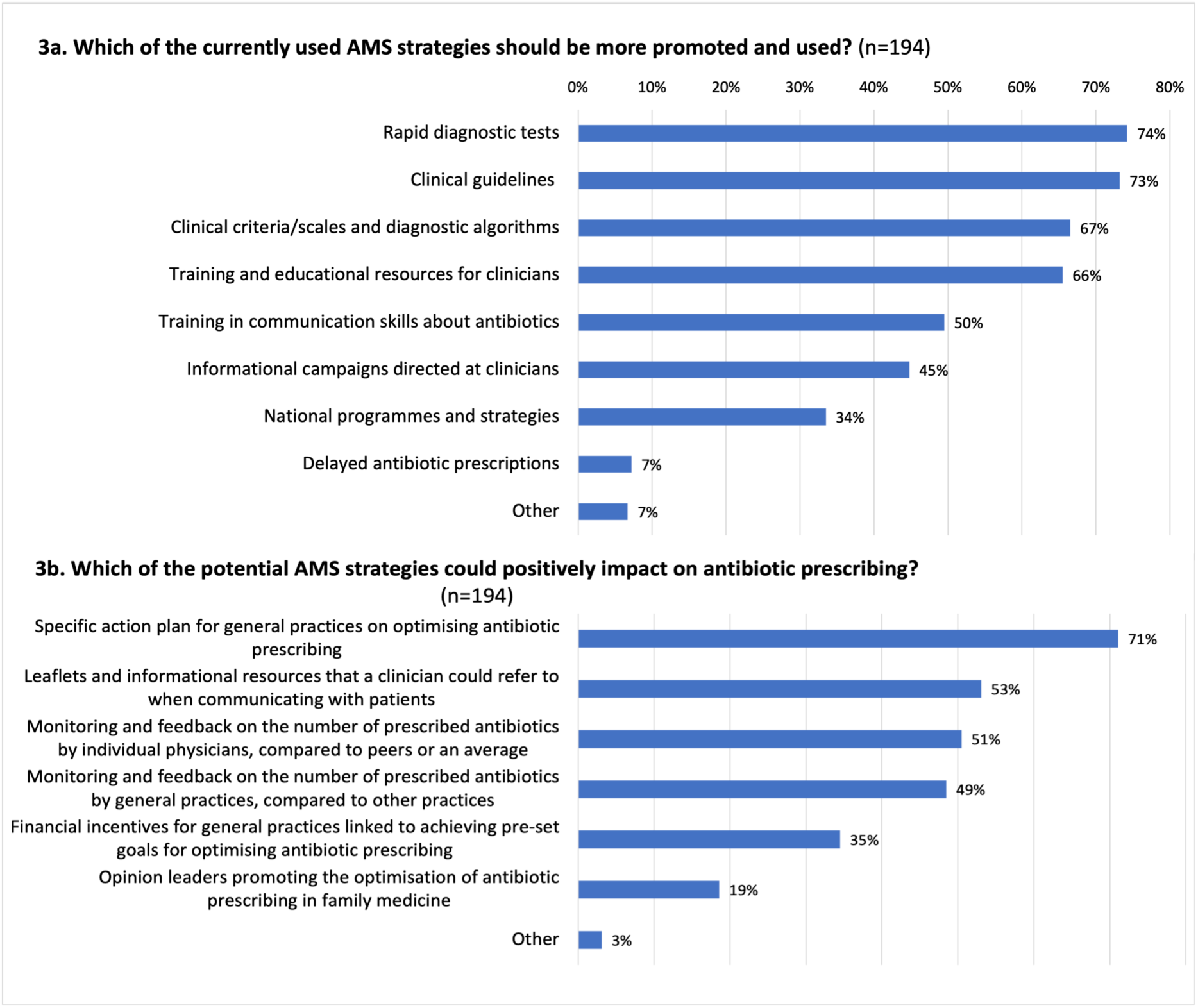
Priority AMS strategies for further promotion and use.

**Figure 4.**
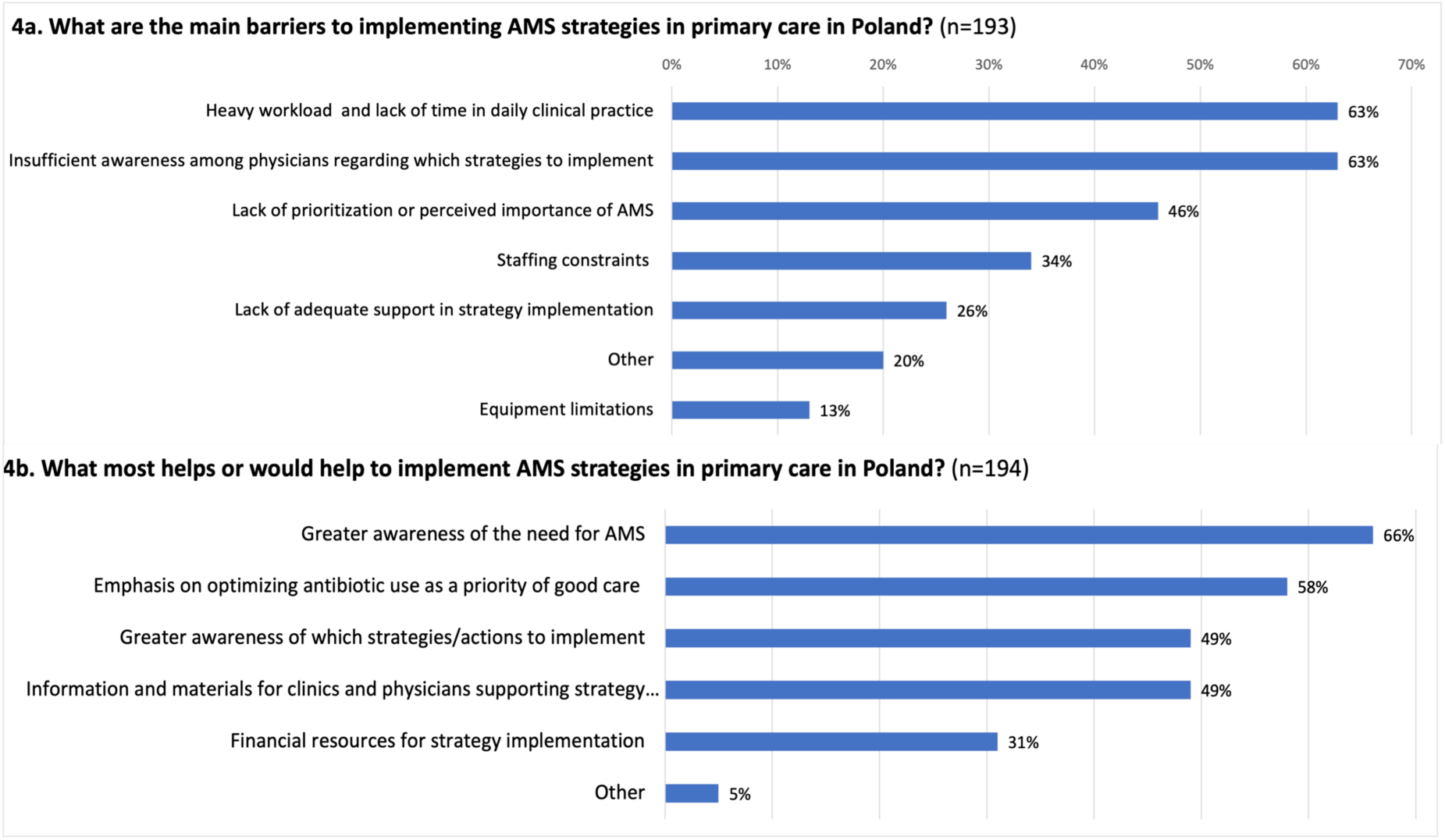
Barriers and facilitators to implementing AMS strategies.

**Table 1.** Characteristics of the study group.

| Characteristic | Category | Number of respondents (N=270) | Proportion of respondents (%) |
|---|---|---|---|
| **Medical specialisation** <sup>a</sup> | Specialist in Family Medicine | 112 | 41.5 |
|  | Resident (In training) in Family Medicine | 72 | 26.7 |
|  | Specialist in Internal Medicine | 42 | 15.6 |
|  | Without specialization | 25 | 9.3 |
|  | Specialist in Paediatrics | 24 | 8.9 |
|  | Other<sup>b</sup> | 7 | 2.6 |
|  | Resident (In training) in Paediatrics | 3 | 1.1 |
| **Work experience in primary care** (range 0.5–45, mean 9.2, median 6.0) | 0.5 to 5 years | 131 | 48.5 |
|  | 6 to 10 years | 64 | 23.7 |
|  | 11 to 15 years | 27 | 10.0 |
|  | 16 to 20 years | 16 | 5.9 |
|  | 21+ years | 32 | 11.9 |
| **Typical working hours in primary care per week** (mean 27.5, median 30)<sup>c</sup> | 3–8 hours (~half to 1 day/week) | 31 | 11.5 |
|  | 9–20 hours (~part-time) | 38 | 14.1 |
|  | 21–40 hours (~full-time) | 182 | 67.4 |
|  | 41 hours and more | 13 | 4.8 |
| **Type of primary care facility** | Publicly funded only | 202 | 74.8 |
|  | Publicly funded and private/commercial | 65 | 24.1 |
|  | Private/commercial only | 3 | 1.1 |
| **Gender** | Female | 209 | 77.4 |
|  | Male | 60 | 22.2 |
|  | Prefer not to say | 1 | 0.4 |
| **Location size of the main primary care facility** <sup>d</sup> | Rural area / village | 39 | 14.4 |
|  | Town (up to 100,000 inhabitants) | 88 | 32.6 |
|  | City (100,000–500,000 inhabitants) | 63 | 23.3 |
|  | Large city (over 500,000 inhabitants) | 80 | 29.6 |

*Note.* <sup>a</sup> Categories are non-exclusive as some respondents indicated more than one category (e.g., had more than one specialisation). <sup>b</sup> Other specialisations included: Nephrology, Neonatology, Palliative Medicine, Allergology, Forensic Medicine, and Resident (In training) in Neonatology. <sup>c</sup> Six responses, indicating 60, 100, 100, 150 and 150 working hours in primary care per week, were treated as missing data. They were considered unfeasible and were probably provided erroneously. <sup>d</sup> Percentages may not sum up to 100.0% due to rounding.

Eligible respondents were physicians (family medicine, internal medicine, and paediatrics specialists, as well as trainees and physicians without a speciality) practising in primary care in Poland for at least 6 months and fluent in Polish. Convenience sampling was used. The survey was advertised via social media in closed groups of physicians verified by their practice licence, newsletters of the Polish Society of Family Medicine (PTMR), professional networks and contacts of the study team, paid social media adverts, and direct emails to 48 general practices randomly selected from all 16 voivodeships/provinces. Data were collected between March and June 2026 via the Qualtrics online platform. Access to the survey was restricted to those with the provided link. Participation was voluntary, anonymous, and without reimbursement. At the start of the survey, respondents received study information and provided informed consent. They could withdraw at any time without providing a reason.

Descriptive analyses were conducted, with percentages calculated as proportions of valid responses for each question/item. Proportions of respondents and the number of responses for each question/item are reported. Free-text comments were categorised by topic and content and reported narratively. Correlational analyses to address the pre-registered secondary objective were not conducted, as they were statistically unjustified given the low response variance.

## Results

Two hundred ninety respondents were eligible and consented to participate, and 270 completed the required questions about their characteristics. Response counts differed because later sections and questions were optional (see flowchart in Supplement 3). Respondents provided 65 optional free-text comments, which we categorised as specific AMS strategies (n=16), barriers/challenges (n=51), and facilitators and suggestions (n=15). Table 1 presents sample characteristics, and Supplement 4 reports respondents’ work locations (rural/urban, voivodeships/provinces).

Most respondents strongly agreed or agreed that AMS is important (93%), that they play an important role in AMS and in preventing AMR (89%), and that AMR is a significant threat in primary care (88%) (Figure 1). Respondents’ familiarity with, use of, confidence in using, and perceived effectiveness of AMS strategies are reported in Figure 2 (also in Supplement 5); the self-reported frequency of use is shown in Table 2.

**Table 2.**
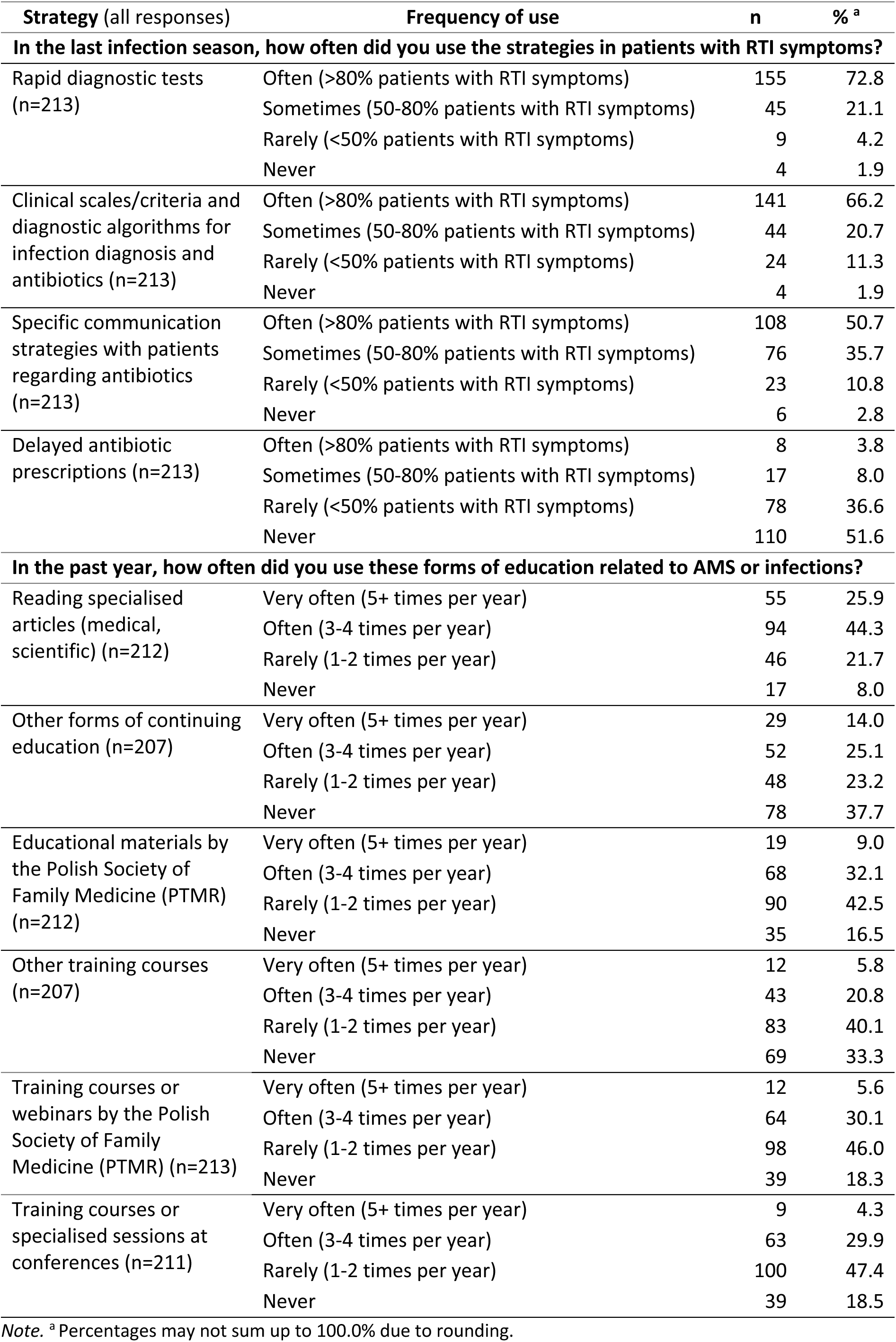
Self-reported frequency of use of AMS strategies.

| **Strategy** (all responses) | **Frequency of use** | **n** | **%** <sup>a</sup> |
|---|---|---|---|
| **In the last infection season, how often did you use the strategies in patients with RTI symptoms?** |  |  |  |
| Rapid diagnostic tests (n=213) | Often (>80% patients with RTI symptoms) | 155 | 72.8 |
|  | Sometimes (50-80% patients with RTI symptoms) | 45 | 21.1 |
|  | Rarely (<50% patients with RTI symptoms) | 9 | 4.2 |
|  | Never | 4 | 1.9 |
| Clinical scales/criteria and diagnostic algorithms for infection diagnosis and antibiotics (n=213) | Often (>80% patients with RTI symptoms) | 141 | 66.2 |
|  | Sometimes (50-80% patients with RTI symptoms) | 44 | 20.7 |
|  | Rarely (<50% patients with RTI symptoms) | 24 | 11.3 |
|  | Never | 4 | 1.9 |
| Specific communication strategies with patients regarding antibiotics (n=213) | Often (>80% patients with RTI symptoms) | 108 | 50.7 |
|  | Sometimes (50-80% patients with RTI symptoms) | 76 | 35.7 |
|  | Rarely (<50% patients with RTI symptoms) | 23 | 10.8 |
|  | Never | 6 | 2.8 |
| Delayed antibiotic prescriptions (n=213) | Often (>80% patients with RTI symptoms) | 8 | 3.8 |
|  | Sometimes (50-80% patients with RTI symptoms) | 17 | 8.0 |
|  | Rarely (<50% patients with RTI symptoms) | 78 | 36.6 |
|  | Never | 110 | 51.6 |
| **In the past year, how often did you use these forms of education related to AMS or infections?** |  |  |  |
| Reading specialised articles (medical, scientific) (n=212) | Very often (5+ times per year) | 55 | 25.9 |
|  | Often (3-4 times per year) | 94 | 44.3 |
|  | Rarely (1-2 times per year) | 46 | 21.7 |
|  | Never | 17 | 8.0 |
| Other forms of continuing education (n=207) | Very often (5+ times per year) | 29 | 14.0 |
|  | Often (3-4 times per year) | 52 | 25.1 |
|  | Rarely (1-2 times per year) | 48 | 23.2 |
|  | Never | 78 | 37.7 |
| Educational materials by the Polish Society of Family Medicine (PTMR) (n=212) | Very often (5+ times per year) | 19 | 9.0 |
|  | Often (3-4 times per year) | 68 | 32.1 |
|  | Rarely (1-2 times per year) | 90 | 42.5 |
|  | Never | 35 | 16.5 |
| Other training courses (n=207) | Very often (5+ times per year) | 12 | 5.8 |
|  | Often (3-4 times per year) | 43 | 20.8 |
|  | Rarely (1-2 times per year) | 83 | 40.1 |
|  | Never | 69 | 33.3 |
| Training courses or webinars by the Polish Society of Family Medicine (PTMR) (n=213) | Very often (5+ times per year) | 12 | 5.6 |
|  | Often (3-4 times per year) | 64 | 30.1 |
|  | Rarely (1-2 times per year) | 98 | 46.0 |
|  | Never | 39 | 18.3 |
| Training courses or specialised sessions at conferences (n=211) | Very often (5+ times per year) | 9 | 4.3 |
|  | Often (3-4 times per year) | 63 | 29.9 |
|  | Rarely (1-2 times per year) | 100 | 47.4 |
|  | Never | 39 | 18.5 |

*Note.* <sup>a</sup> Percentages may not sum up to 100.0% due to rounding.

### Rapid diagnostic tests, clinical guidelines and scales/criteria

Nearly all respondents partially or fully agreed that they were familiar with POCTs (99.5%, n=211), regularly used POCTs (93.3%, n=205), were confident in using POCTs (97.7%, n=208), and considered POCTs effective in AMS (98.1%, n=207). 93.9% (n=200) reported using POCTs often (with >80% of patients with RTI) or sometimes (with 50-80% of patients with RTI) during the last infection season. Similarly, nearly all respondents were at least partly familiar with clinical guidelines (99.1%, n=211) and clinical scales/criteria/algorithms (98.1%, n=209), regularly referred to guidelines (89.2%, n=190), used clinical scales (83.9%, n=177), and considered guidelines (95.3%, n=202) and scales (95.3%, n=202) effective. 94.3% (n=199) were confident in using clinical scales, and 86.9% (n=185) used them often or sometimes with RTI patients.

### Communication strategies with patients

Nearly all respondents partially or fully agreed that they were familiar with communication strategies with patients about antibiotics (95.8%, n=204), regularly used them (86.8%, n=184), felt confident using them (93.9%, n=200), and thought these strategies were effective (92.5%, n=196). However, only 50.7% (n=108) reported using them with >80% of RTI patients.

### National-level strategies: AMR actions, informational campaigns, education and training

83.1% (n=177) of respondents partly or fully agreed that they were familiar with national AMR actions, and 80.2% (n=170) considered them (partly) effective. However, only 55.7% (n=118) regularly referred to such action plans. Informational/educational campaigns were known to only 64.3% (n=137) of respondents, and only 57.6% (n=122) considered them effective.

Only 54.5% (n=116) of respondents reported regularly participating in education and training related to antibiotics, AMR or infections, with reading specialised articles the most common form of education (70.3%, n=149). A minority of respondents often or very often engaged with educational materials/resources from the Polish Society of Family Medicine (PTMR) (41.0%, n=87), other forms of continuing education (39.1%, n=81), PTMR training courses/webinars (35.7%, n=76), or specialised training sessions at conferences (34.1%, n=72). Nevertheless, 91.5% (n=194) of respondents (partially) agreed that education/training is effective, and 87.8% (n=187) that it increased their knowledge.

### Delayed antibiotic prescriptions

Although 74.9% (n=158) of respondents agreed or partially agreed that they were familiar with delayed antibiotic prescriptions, only 15.3% (n=32) regularly used them, and only 11.8% (n=25) used them often or sometimes during the last infection season. 47.6% (n=98) were (partially) confident in using delayed prescriptions, and 50.0% (n=105) (partially) agreed that they were effective in AMS. Free-text comments reiterated that physicians did not use delayed prescriptions, with one respondent calling it an “anti-strategy”. Comments described a lack of trust that patients would use delayed prescriptions appropriately, a belief that they would use antibiotics immediately or in the future without consultation, and a view that patients cannot decide when to use antibiotics. Respondents reported preferring to prescribe immediate antibiotics or advise follow-up visits. However, three respondents suggested circumstances in which delayed prescriptions could or were used: with engaged patients or before periods of limited healthcare access (e.g., a holiday).

### Suggestions for implementation

Among the key strategies respondents thought should be promoted and used more were those already most familiar and used: rapid tests, clinical guidelines, clinical criteria/scales, and training and educational resources (Figure 3a). AMS action plans for general practices were the top potential strategy (i.e. not yet implemented but one that could have a positive impact; Figure 3b). Of 19 free-text suggestions, 52.6% (n=10) called for patient education. Other suggestions included rapid CRP POCTs; monitoring (auditing) antibiotic prescribing in randomly selected general practices and in out-of-hours (where antibiotic prescribing was described as often not clinically indicated); a verifiable obligation for continuing education; more accessible resources/training (e.g., short videos, podcasts); and encouraging older clinicians to reflect on their prescribing.

### Perceived barriers and facilitators to implementation

As the main barriers and challenges to implementing AMS (Figure 4a), respondents considered a lack of time in daily practice and insufficient awareness among physicians of which strategies to implement. As the main facilitators (Figure 4b), respondents indicated greater awareness of the need for AMS and emphasised antibiotic optimisation as a priority in good healthcare.

Fifty-one free-text comments described perceived barriers and challenges to AMS and appropriate antibiotic use. Twenty-five comments highlighted patient-related factors, including patient expectations, demand and pressure for antibiotics; patient/public beliefs about the effectiveness and necessity of antibiotics; and a lack of patient/public knowledge, awareness and perceived relevance in this area. Patients were described as misusing antibiotics (e.g., starting or obtaining antibiotics before consulting physicians), behaving “aggressively” when demanding antibiotics, and slandering doctors online if they refused antibiotics. Consequently, suggestions focused on patient and school education, and on patient/public leaflets and campaigns.

Other comments attributed inappropriate antibiotic prescribing to “other” physicians, such as “older” physicians, those without a specialisation, laryngologists, paediatricians, or “random” physicians working in primary care. Inappropriate prescribing was attributed to unfamiliarity with clinical guidelines, belief that antibiotics are “good for everything”, prescribing without a clinical indication, or failing to differentiate between viral and bacterial aetiology. It was also linked to interactions with patients: physicians’ lack of “assertiveness” and prescribing antibiotics to avoid confrontation or reconsultations. These beliefs and behaviours were sometimes linked to perceived difficult-to-change habits, expectations, and cultural norms. Corresponding suggestions included evidence that overuse of antibiotics does not lead to better therapeutic outcomes, “standardised schemas for what, when, in which infection” to prescribe, limiting the use of topical antibiotics, ensuring awareness and compliance with clinical guidelines, improving access to physicians, providing initial health advice in pharmacies, and removing the requirement for sick leave certificates.

## Discussion

Overall, surveyed physicians believed in the threat of AMR, the importance of, and their role in, AMS. Rapid diagnostic tests, clinical guidelines, and clinical scales/criteria were the most recognised, utilised, considered effective, and recommended strategies. Contrastingly, delayed prescriptions were the least used, perceived as least effective, and least recommended. Insufficient awareness of which strategies to implement (a key barrier) corresponded with the suggested increasing awareness of the need for AMS and emphasising it as a priority of good healthcare (a key facilitator). Table 3 summarises the implications for practice and research.

**Table 3.** Key implications for practice and research.

| **Strategies & Suggestions** | **Implications for practice** | **Implications for research** |
|---|---|---|
| Awareness of the importance of AMR/AMS and physicians’ role in them in primary care | - Target specific sub-groups of physicians to raise awareness of AMR risks and AMS strategies in primary care.<br>- Increase awareness of which AMS strategies to implement and how.<br>- Emphasise AMS/antibiotic optimisation as a priority of good care. |  |
| Rapid diagnostic tests | Incorporate diagnostic tests with other, complementary strategies (e.g., guidelines, communication skills). | Evaluate the impact of test use on antibiotic prescribing in routine care. |
| Clinical guidelines and clinical scales/criteria/algorithms | Increase adherence to clinical guidelines and scales/criteria among physicians who do not use them regularly. |  |
| Communication strategies about antibiotics/infections | - Increase the use of specific communication strategies to address challenges in physician–patient interaction (e.g. addressing patient demand for antibiotics).<br>- Make available resources (e.g. leaflets) to support communication. | Evaluate enhanced communication skills training in routine care. |
| Tasks/strategies within a national action plan | - Promote AMS actions/strategies specific to primary care.<br>- Facilitate feedback on and peer comparisons of antibiotic prescribing. | Evaluate the implementation and impact of tasks/ strategies included in a national action plan. |
| Informational/ educational campaigns | Increase the reach of such campaigns. | Evaluate the reach and effectiveness of campaigns. |
| Clinician-targeted education and training | - Increase and monitor the engagement of all physicians in regular education/ training related to AMS/AMR.<br>- Identify and develop most needed, acceptable, and effective education strategies. | Evaluate the implementation and effectiveness of different forms of clinician education/training. |

Rapid diagnostic testing was identified as a key strategy for optimising antibiotic prescribing, a finding consistent with recent surveys in Poland [18,20]. This aligns with the evidence [3–5]and international recommendations that antibiotic prescribing in primary care should be guided by diagnostics [24]. However, a recent large randomised controlled trial in European (including Polish) primary care found that POCTs for RTI did not reduce antibiotic prescribing, concluding that multiple strategies are needed to address clinical and non-clinical factors influencing prescribing [25,26]. Similarly, implementing diagnostics in routine English care was not associated with lower prescribing [21]. In 2022, the National Health Service in Poland began funding selected POCTs; their use and impact on antibiotic prescribing should be evaluated.

Despite high familiarity with, confidence in using, and perceived effectiveness of communication strategies, and reported challenges with patient expectations/demand for antibiotics, only 51% of respondents reported using communication strategies often, 50% recommended communication-related training, and 53% recommended resources to support communication. Specific communication strategies could help identify and address patient expectations and provide information effectively, particularly because research shows that physicians tend to overestimate patients’ expectations for antibiotics [18]. Communication skills training combined with POCTs was found to be more effective, and to have a longer-lasting impact, than POCTs or communication alone [27,28]. Patient leaflets used interactively to support communication about infections and antibiotics also have encouraging effects [3,5,27], and were recommended by our respondents. Communication training on strategies that fit within the time constraints of general practice might address patient-related drivers of inappropriate antibiotic prescribing and strengthen other strategies, such as adherence to clinical guidelines and POCTs.

The EU recommends strengthening national AMS programmes [1,24]. The Polish National Health Programme [16] lacks specific tasks or targets for primary care. Our results suggest that implementing the national approach could be strengthened by raising awareness of which AMS strategies to implement and how, supporting the development of primary care AMS action plans, and facilitating feedback on antibiotic prescribing and peer comparisons. Such strategies, promoted nationally and operationalised locally, helped reduce antibiotic use in England [21,22]. Because uptake and sustained implementation of AMS strategies in routine care are challenging [12,13], support is needed at multiple system levels to advance AMS in Poland.

Finally, we found high awareness of AMR risks and the need for AMS. Others found that physician awareness and knowledge of antibiotics and AMR are high, yet they recommended more education [17–20][17–20]. While knowledge is critical, it alone is insufficient for behaviour change, particularly for complex behaviours such as antibiotic prescribing. Some physicians may benefit from education, given that 5–8% of our respondents still disagreed with the risk of AMR and the importance of AMS and their role in it. However, more physicians might benefit from enhancing their motivation by encouraging them to prioritise prudent antibiotic prescribing as part of good-quality care and by addressing other (clinical and non-clinical) factors influencing antibiotic prescribing [12,20,26].

### Strengths and limitations

The study was pre-registered and conducted by a multidisciplinary team. We established the questionnaire’s face validity through piloting and review by the target respondents. We recruited a diverse sample that broadly resembled national patterns (e.g., high proportions of family medicine specialists, women, and those working in publicly funded clinics, Supplement 4). However, the sample was non-representative, and convenience sampling may have led to over-recruitment of female and younger physicians, those more interested in research and/or the topic, and more active social media users. Familiarity with and use of AMS strategies might be higher among respondents than among physicians who were not reached or declined to participate. Future research should validate the results with a representative sample. While our findings offer valuable insights in a country with high antibiotic consumption, external validity to countries with different healthcare structures, antibiotic prescribing rates, or stewardship policies may be limited. Because our recruitment was open and the total number of eligible physicians exposed to the advertisements is unknown, we cannot calculate a response rate. Instead, we have transparently reported response numbers.

## Conclusion

Physicians working in Polish primary care report high awareness of the threat of AMR, the importance of AMS and their role in it, and familiarity with and use of evidence-based AMS strategies. However, a minority of physicians remain less (or not) concerned about AMR/AMS. To address barriers and support AMS progress, key recommendations include improving physicians’ diagnostic precision, developing specific AMS action plans that fit within high-workload settings, supporting communication and addressing patient demand for antibiotics, and promoting awareness of the need for AMS and its role in high-quality care. Evidence is needed on the impact of AMS strategies on antibiotic prescribing in routine care.

## Data Availability

All data produced in the present study will be deposited in the SWPS Science Share repository and are available upon reasonable request to the authors. Data tables are provided in Supplemental Materials.

## Declarations

### Ethical approval

The study was reviewed and approved by the institutional Research Ethics Committee at the Department of Psychology in Wrocław, SWPS University; ethics approval reference: 01/E/03/2026. The study was conducted in line with the Declaration of Helsinki. Respondents gave informed consent before entering the anonymous online survey.

### Author contributions

AJB: Conceptualisation, Data curation, Formal analysis, Funding acquisition, Investigation, Methodology, Project administration, Visualisation, Writing – original draft. IPP: Conceptualisation, Methodology, Writing – review & editing. JK: Conceptualisation, Methodology, Writing – review & editing. AL: Conceptualisation, Methodology, Writing – review & editing. KK: Conceptualisation, Methodology, Writing – review & editing. MB: Conceptualisation, Methodology, Writing – review & editing. All authors approved the manuscript for publication.

### Declaration of generative AI use

Generative AI (Gemini, version 3.6 Flash) was used to translate the questionnaire from Polish (original) into English. The first author, AJB, checked and edited the translation and takes responsibility for its accuracy. AI-supported software (Grammarly) was used to check and correct the text originally written by the authors, who take responsibility for the text’s accuracy.

### Data availability statement

The data tables generated during this study and supporting the results are provided in Supplemental Materials. The dataset will be deposited and made available in the SWPS Science Share repository.

## Supplemental materials

Supplement 1: Reporting checklist

Supplement 2: The questionnaire

Supplement 3: Flow diagram of the number of respondents

Supplement 4: Additional respondent characteristics Supplement 5: Data tables

