## Supplemental Materials for "Antimicrobial stewardship strategies to optimise antibiotic prescribing in primary care: physicians’ awareness, beliefs, and strategy use"

### Supplement 1: Reporting checklist

#### Table S1. Checklist for Reporting Of Survey Studies (CROSS)

Sharma A, Minh Duc NT, Luu Lam Thang T, et al. A Consensus-Based Checklist for Reporting of Survey Studies (CROSS). J GEN INTERN MED. 2021;36(10):3179–3187.

| Section/topic | Item | Item description | Reported on page # or in supplement (S) # |
|---|---|---|---|
| **Title and abstract** | | | |
| | 1a | State the word “survey” along with a commonly used term in title or abstract to introduce the study’s design. | 2 |
| Title and abstract | 1b | Provide an informative summary in the abstract, covering background, objectives, methods, findings/results, interpretation/discussion, and conclusions. | 2 |
| **Introduction** | | | |
| Background | 2 | Provide a background about the rationale of study, what has been previously done, and why this survey is needed. | 3 |
| Purpose/aim | 3 | Identify specific purposes, aims, goals, or objectives of the study. | 3 |
| **Methods** | | | |
| Study design | 4 | Specify the study design in the methods section with a commonly used term (e.g., cross-sectional or longitudinal). | 3 |
| | 5a | Describe the questionnaire (e.g., number of sections, number of questions, number and names of instruments used). | 4, Suppl. 2 |
| | 5b | Describe all questionnaire instruments that were used in the survey to measure particular concepts. Report target population, reported validity and reliability information, scoring/classification procedure, and reference links (if any). | 4, Suppl. 2 |
| Data collection methods | 5c | Provide information on pretesting of the questionnaire, if performed (in the article or in an online supplement). Report the method of pretesting, number of times questionnaire was pre-tested, number and demographics of participants used for pretesting, and the level of similarity of demographics between pre-testing participants and sample population. | 4 |
| | 5d | Questionnaire if possible, should be fully provided (in the article, or as appendices or as an online supplement). | Suppl. 2 |
| | 6a | Describe the study population (i.e., background, locations, eligibility criteria for participant inclusion in survey, exclusion criteria). | 4 |
| Sample characteristics | 6b | Describe the sampling techniques used (e.g., single stage or multistage sampling, simple random sampling, stratified sampling, cluster sampling, convenience sampling). Specify the locations of sample participants whenever clustered sampling was applied. | 4 |
| | 6c | Provide information on sample size, along with details of sample size calculation. | N/A – Target sample size was reported in the pre-registration, |

| | | | |
|---|---|---|---|
| | | | but it was calculated for correlational analyses which were not conducted |
| | 6d | Describe how representative the sample is of the study population (or target population if possible), particularly for population-based surveys. | Suppl. 4, 8 |
| Survey administration | 7a | Provide information on modes of questionnaire administration, including the type and number of contacts, the location where the survey was conducted (e.g., outpatient room or by use of online tools, such as SurveyMonkey). | 4 |
| | 7b | Provide information of survey's time frame, such as periods of recruitment, exposure, and follow-up days. | 4 |
| | 7c | Provide information on the entry process:<br>–>For non-web-based surveys, provide approaches to minimize human error in data entry.<br>–>For web-based surveys, provide approaches to prevent "multiple participation" of participants. | 4<br>4<br>(participation with the link only, participants could complete unfinished survey later, rather than participating multiple times) |
| Study preparation | 8 | Describe any preparation process before conducting the survey (e.g., interviewers' training process, advertising the survey). | 4 |
| Ethical considerations | 9a | Provide information on ethical approval for the survey if obtained, including informed consent, institutional review board [IRB] approval, Helsinki declaration, and good clinical practice [GCP] declaration (as appropriate). | 4, 8 |
| | 9b | Provide information about survey anonymity and confidentiality and describe what mechanisms were used to protect unauthorized access. | 4 |
| Statistical analysis | 10a | Describe statistical methods and analytical approach. Report the statistical software that was used for data analysis. | 4 |
| | 10b | Report any modification of variables used in the analysis, along with reference (if available). | N/A |
| | 10c | Report details about how missing data was handled. Include rate of missing items, missing data mechanism (i.e., missing completely at random [MCAR], missing at random [MAR] or missing not at random [MNAR]) and methods used to deal with missing data (e.g., multiple imputation). | N/A (only the collected responses were analysed) |
| | 10d | State how non-response error was addressed. | N/A |
| | 10e | For longitudinal surveys, state how loss to follow-up was addressed. | N/A |
| | 10f | Indicate whether any methods such as weighting of items or propensity scores | N/A |

| | | | |
|---|---|---|---|
| | | have been used to adjust for non-representativeness of the sample. | |
| | 10g | Describe any sensitivity analysis conducted. | N/A |
| **Results** | | | |
| Respondent characteristics | 11a | Report numbers of individuals at each stage of the study. Consider using a flow diagram, if possible. | 4, Suppl. 3 |
| | 11b | Provide reasons for non-participation at each stage, if possible. | Unknown |
| | 11c | Report response rate, present the definition of response rate or the formula used to calculate response rate. | 8 |
| | 11d | Provide information to define how unique visitors are determined. Report number of unique visitors along with relevant proportions (e.g., view proportion, participation proportion, completion proportion). | Data not available |
| Descriptive results | 12 | Provide characteristics of study participants, as well as information on potential confounders and assessed outcomes. | Table 1, Suppl. 4 |
| Main findings | 13a | Give unadjusted estimates and, if applicable, confounder-adjusted estimates along with 95% confidence intervals and p-values. | N/A |
| | 13b | For multivariable analysis, provide information on the model building process, model fit statistics, and model assumptions (as appropriate). | N/A |
| | 13c | Provide details about any sensitivity analysis performed. If there are considerable amount of missing data, report sensitivity analyses comparing the results of complete cases with that of the imputed dataset (if possible). | N/A |
| **Discussion** | | | |
| Limitations | 14 | Discuss the limitations of the study, considering sources of potential biases and imprecisions, such as non-representativeness of sample, study design, important uncontrolled confounders. | 8 |
| Interpretations | 15 | Give a cautious overall interpretation of results, based on potential biases and imprecisions and suggest areas for future research. | 8 |
| Generalizability | 16 | Discuss the external validity of the results. | 8 |
| **Other sections** | | | |
| Role of funding source | 17 | State whether any funding organization has had any roles in the survey's design, implementation, and analysis. | 8 |
| Conflict of interest | 18 | Declare any potential conflict of interest. | 8 |
| Acknowledgements | 19 | Provide names of organizations/persons that are acknowledged along with their contribution to the research. | N/A |

### Supplement 2: The questionnaire used in the study

#### Introduction

#### Section 1. Screening and consent

*[Required questions; only those answering "yes" to both questions were allowed to proceed to the survey]*

| | |
|---|---|
| Czy potwierdza Pan/Pani zapoznanie się z informacjami o badaniu oraz wyraża Pan/Pani zgodę na udział w badaniu? [tak, nie] | Do you confirm that you have read and understood the information about the study, and do you consent to participate in it? [yes, no] |
| Czy potwierdza Pan/Pani, że pracuje jako lekarz w placówce POZ/lekarza rodzinnego od co najmniej 6 miesięcy? [tak, nie] | Do you confirm that you have been working as a physician in a primary care facility for at least 6 months? [yes, no] |

#### Section 2. Respondent characteristics

*[All questions in this section were required]*

| Question | Polish version (original) | English version (translation) |
|---|---|---|
| **Q1** | **Jaka jest Pana/Pani specjalizacja medyczna?** *(Pytanie wielokrotnego wyboru)*<br><br>a. Bez specjalizacji<br>b. Specjalista medycyny rodzinnej<br>c. Specjalista pediatrii<br>d. Specjalista chorób wewnętrznych<br>e. W trakcie specjalizacji z medycyny rodzinnej<br>f. W trakcie specjalizacji z pediatrii<br>g. W trakcie specjalizacji z chorób wewnętrznych<br>h. Inna - Jaka? | **What is your medical specialty?** *(Select all that apply)*<br><br>a. None / Non-specialist<br>b. Family Medicine Specialist<br>c. Paediatric Specialist<br>d. Internal Medicine Specialist<br>e. Resident in Family Medicine<br>f. Resident in Paediatrics<br>g. Resident in Internal Medicine<br>h. Other – Please specify |
| **Q2** | **Proszę podać staż pracy (w latach) w placówce/placówkach POZ.** *(Pytanie otwarte)* | **Please state your years of experience in primary care facility/facilities.** *(Open-ended question)* |
| **Q3** | **Ile godzin w tygodniu zazwyczaj pracuje Pan/Pani w placówce/placówkach POZ?** (*Pytanie otwarte)* | **On average, how many hours per week do you work in primary care facility/facilities?** *(Open-ended question)* |

| | | |
|---|---|---|
| **Q4** | **W jakim typie lub typach placówki POZ Pan/Pani pracuje?** *(Proszę wybrać wszystkie, które się odnoszą.)*<br><br>a. Świadczących usługi tylko w ramach NFZ<br>b. Świadczących usługi w ramach NFZ oraz komercyjnie (np. LuxMed)<br>c. Świadczących usługi tylko komercyjnie/prywatnie | **In what type(s) of primary care facility do you work?** *(Select all that apply)*<br><br>a. Facilities providing services exclusively under the public healthcare system (National Health Fund (NFZ))<br>b. Facilities providing services both under the public healthcare system and commercially / privately<br>c. Facilities providing services exclusively commercially / privately |
| **Q5** | **W którym województwie Pan/Pani pracuje?** *(Jedna odpowiedź z listy)*<br><br>dolnośląskie, kujawsko-pomorskie, lubelskie, lubuskie, łódzkie, małopolskie, mazowieckie, opolskie, podkarpackie, podlaskie, pomorskie, śląskie, świętokrzyskie, warmińsko-mazurskie, wielkopolskie, zachodniopomorskie | **In which voivodeship (province) do you work?** *(Select one from the list)*<br><br>Lower Silesian, Kuyavian-Pomeranian, Lublin, Lubusz, Łódź, Lesser Poland, Masovian, Opole, Subcarpathian, Podlaskie, Pomeranian, Silesian, Holy Cross, Warmian-Masurian, Greater Poland, West Pomeranian |
| **Q6** | **Jakiej wielkości jest miejscowość, w której Pan/Pani obecnie głównie pracuje?** (Proszę wybrać jedną opcję, odnoszącą się do miejsca, w którym Pan/Pani najwięcej lub najczęściej pracuje.)<br><br>a. Wieś<br>b. Małe miasto do 100 tys. mieszkańców<br>c. Średnie miasto z 100-500 tys. mieszkańców<br>d. Duże miasto powyżej 500 tys. Mieszkańców | **What is the size of the locality/city where you currently work most of your time?** (Please select the single option that corresponds to the location where you work the most or most frequently.)<br><br>a. Rural area / Village<br>b. Small town / city (up to 100,000 inhabitants)<br>c. Medium-sized city (100,000–500,000 inhabitants)<br>d. Large city (over 500,000 inhabitants) |
| **Q7** | **Proszę podać swoją płeć.**<br><br>a. Kobieta<br>b. Mężczyzna<br>c. Inna<br>d. Wolę nie podawać | **Please specify your gender.**<br><br>a. Female<br>b. Male<br>c. Other<br>d. Prefer not to say |

#### Section 3. Perceived role in and importance of antimicrobial stewardship (AMS)

W kolejnych pytaniach odnosimy się do **"ochrony antybiotyków",** którą definiujemy jako **działania mające na celu optymalizację stosowania antybiotyków i minimalizowania niezamierzonych**

**skutków**, takich jak rozwój lekooporności. W praktyce oznacza to dążenie do stosowania właściwego leku, w odpowiedniej dawce, drogą o optymalnej biodostępności oraz przez optymalny czas.

In the following questions, we refer to "antibiotic stewardship," which we define as actions aimed at optimizing antibiotic use and minimizing unintended consequences, such as the development of drug resistance. In practice, this means striving to use the right drug, at the correct dose, via the optimal route of administration, and for the optimal duration.

| Question | Polish version (original) | English version (translation) |
|---|---|---|
| **Q8** | **Jako lekarz pracujący w placówce POZ, w jakim stopniu zgadza się Pan/Pani z poniższymi stwierdzeniami?**<br>*(Skala: 1 - zdecydowanie się nie zgadzam, 2 - nie zgadzam się, 3 - ani się nie zgadzam ani się zgadzam, 4 - zgadzam się, 5 - zdecydowanie się zgadzam)*<br><br>a. Odgrywam istotną rolę w ochronie antybiotyków przed rozwojem lekooporności.<br><br>b. Ochrona i optymalne stosowanie antybiotyków są ważnym elementem opieki nad pacjentami w podstawowej opiece zdrowotnej.<br><br>c. Antybiotykooporność jest istotnym zagrożeniem dla pacjentów w podstawowej opiece zdrowotnej. | **As a primary care physician, to what extent do you agree or disagree with the following statements?**<br>*(Scale: 1 – Strongly disagree, 2 – Disagree, 3 – Neither agree nor disagree, 4 – Agree, 5 – Strongly agree)*<br><br>a. I play an important role in antibiotic stewardship to prevent the development of drug resistance.<br><br>b. Antibiotic stewardship and the optimal use of antibiotics are an important element of patient care in primary care.<br><br>c. Antimicrobial resistance is a significant threat to patients in primary care. |

#### Section 4. Views on and experiences of AMS strategies

W tej sekcji pytamy o Pana/Pani doświadczenia i poglądy związane z **przykładowymi działaniami** w celu ochrony i optymalizacji stosowania antybiotyków i przeciwdziałania antybiotykooporności.

In this section, we ask about your experiences and views regarding example actions aimed at antibiotic stewardship, optimising antibiotic use, and combating antimicrobial resistance.

| Question | Polish version (original) | English version (translation) |
|---|---|---|
| **Q9** | **Czy zgadza się Pan/Pani z zaprezentowanymi stwierdzeniami w odniesieniu do poniższych typów strategii?**<br>*(Opcje odpowiedzi: Tak, Nie, Częściowo, Nie dotyczy)* | **Do you agree with the following statements regarding these types of strategies?**<br>*(Response options: Yes / No / Partially / Not applicable)* |

| | | |
|---|---|---|
| | **Działania w zakresie oporności na środki przeciwdrobnoustrojowe w ramach krajowych programów i planów** (np. Narodowego Programu Zdrowia, Narodowego Programu Ochrony Antybiotyków).<br><br>a. Znam ten typ strategii.<br>b. Regularnie odnoszę się do tego typu strategii.<br>c. Uważam, że ten typ strategii jest efektywny w ochronie antybiotyków. | **Actions addressing antimicrobial resistance within national programs and plans** (e.g., the National Health Program, National Antibiotic Protection Program).<br><br>a. I am familiar with this type of strategy.<br>b. I regularly refer to this type of strategy.<br>c. I believe this type of strategy is effective in antibiotic stewardship. |
| **Q10** | **Czy zgadza się Pan/Pani z zaprezentowanymi stwierdzeniami w odniesieniu do poniższych typów strategii?**<br>*(Opcje odpowiedzi: Tak, Nie, Częściowo, Nie dotyczy)*<br><br>**Wytyczne i rekomendacje kliniczne związane z leczeniem infekcji i stosowaniem antybiotykoterapii w medycynie rodzinnej** (np. Antybiotykoterapia Praktyczna Prof. Dzierżanowskiej, wytyczne i rekomendacje diagnostyki i leczenia określonych chorób zakaźnych PTMR).<br><br>a. Znam ten typ strategii.<br>b. Regularnie odnoszę się do tego typu strategii.<br>c. Uważam, że ten typ strategii jest efektywny w ochronie antybiotyków. | **Do you agree with the following statements regarding these types of strategies?**<br>*(Response options: Yes / No / Partially / Not applicable)*<br><br>**Clinical guidelines and recommendations related to the treatment of infections and antibiotic therapy in family medicine** (e.g., *Practical Antibiotic Therapy* by Prof. Dzierżanowska, guidelines and recommendations for the diagnosis and treatment of specific infectious diseases by the Polish Society of Family Medicine / PTMR).<br><br>a. I am familiar with this type of strategy.<br>b. I regularly refer to this type of strategy.<br>c. I believe this type of strategy is effective in antibiotic stewardship. |
| **Q11** | **Czy zgadza się Pan/Pani z zaprezentowanymi stwierdzeniami w odniesieniu do poniższych typów strategii?**<br>*(Opcje odpowiedzi: Tak, Nie, Częściowo, Nie dotyczy)*<br><br>**Kampanie informacyjne/edukacyjne związane z antybiotykami, antybiotykoopornością lub leczeniem infekcji** (np. Europejski Dzień Wiedzy o Antybiotykach, Światowy Tydzień Wiedzy o Antybiotykooporności).<br><br>a. Znam ten typ strategii. | **Do you agree with the following statements regarding these types of strategies?**<br>*(Response options: Yes / No / Partially / Not applicable)*<br><br>**Information and educational campaigns related to antibiotics, antimicrobial resistance, or the treatment of infections** (e.g., European Antibiotic Awareness Day, World Antimicrobial Awareness Week).<br><br>a. I am familiar with this type of strategy.<br>b. I believe this type of strategy is effective in antibiotic stewardship. |

| | | |
|---|---|---|
| | b. Uważam, że ten typ strategii jest efektywny w ochronie antybiotyków. | |
| **Q12** | **Czy zgadza się Pan/Pani z zaprezentowanymi stwierdzeniami w odniesieniu do poniższych typów strategii?**<br>*(Opcje odpowiedzi: Tak, Nie, Częściowo, Nie dotyczy)*<br><br>**Edukacja i szkolenia związane z antybiotykoterapią, lekoopornością i chorobami zakaźnymi** (np. kursy i materiały edukacyjne PTMR, sesje/szkolenia na konferencjach).<br><br>a. Regularnie biorę udział w takich szkoleniach.<br>b. Takie szkolenia zwiększyły moją wiedzę i umiejętności.<br>c. Uważam, że ten typ strategii jest efektywny w ochronie antybiotyków. | **Do you agree with the following statements regarding these types of strategies?**<br>*(Response options: Yes / No / Partially / Not applicable)*<br><br>**Education and training related to antibiotic therapy, drug resistance, and infectious diseases** (e.g., PTMR courses and educational materials, sessions/training at conferences).<br><br>a. I regularly participate in such training.<br>b. Such training has increased my knowledge and skills.<br>c. I believe this type of strategy is effective in antibiotic stewardship. |
| **Q12a** | **W ostatnim roku, jak często brał/a Pan/Pani udział w poniższych formach dokształcania związanych z ochroną antybiotyków lub chorobami zakaźnymi?**<br>(*Opcje odpowiedzi: nigdy, rzadko [1-2 razy], często [3-4 razy], bardzo często [5+ razy])*<br><br>a. Szkolenia, webinary PTMR<br>b. Szkolenia lub specjalistyczne sesje na konferencjach<br>c. Inne szkolenia | **In the past year, how often did you participate in the following forms of continuing education related to antibiotic stewardship or infectious diseases?**<br>*(Response options: Never / Rarely [1–2 times] / Often [3–4 times] / Very often [5+ times])*<br><br>a. Training courses or webinars by the Polish Society of Family Medicine (PTMR)<br>b. Training courses or specialized sessions at conferences<br>c. Other training courses |
| **Q12b** | **W ostatnim roku, jak często korzystał/a Pan/Pani z poniższych form dokształcania związanych z ochroną antybiotyków lub chorobami zakaźnymi?**<br>*(Opcje odpowiedzi: nigdy, rzadko (1-2 razy), często (3-4 razy), bardzo często (5+ razy))*<br><br>a. Materiały edukacyjne PTMR<br>b. Czytanie specjalistycznych artykułów (medycznych, naukowych)<br>a. Inne formy dokształcania | **In the past year, how often did you use the following forms of continuing education related to antibiotic stewardship or infectious diseases?**<br>*(Response options: Never / Rarely [1–2 times] / Often [3–4 times] / Very often [5+ times])*<br><br>a. Educational materials by the Polish Society of Family Medicine (PTMR)<br>b. Reading specialized articles (medical, scientific) |

| | | |
|---|---|---|
| | | c. Other forms of continuing education |
| **Q12c** | **Jeśli wybrał/a Pan/Pani "inne" formy szkoleń i dokształcania, to proszę napisać jakie.**<br>*(Pytanie otwarte)* | **If you selected "other" training courses or forms of continuing education, please specify:**<br>*(Open-ended question)* |
| **Q13** | **Czy zgadza się Pan/Pani z zaprezentowanymi stwierdzeniami w odniesieniu do poniższych typów strategii?**<br><br>**Skale/kryteria kliniczne i algorytmy diagnostyczne związane z diagnostyką infekcji i antybiotykoterapią** (np. skala Centora).<br><br>a. Znam ten typ strategii.<br>b. Regularnie stosuję tego typu strategie.<br>c. Czuję się pewnie w stosowaniu tego typu strategii.<br>d. Uważam, że ten typ strategii jest efektywny w ochronie antybiotyków. | **Do you agree with the following statements regarding these types of strategies?**<br><br>**Clinical scales/criteria and diagnostic algorithms related to infection diagnosis and antibiotic therapy** (e.g., the Centor score).<br><br>a. I am familiar with this type of strategy.<br>b. I regularly use this type of strategy.<br>c. I feel confident using this type of strategy.<br>d. I believe this type of strategy is effective in antibiotic stewardship. |
| **Q14** | **Czy zgadza się Pan/Pani z zaprezentowanymi stwierdzeniami w odniesieniu do poniższych typów strategii?**<br><br>**Szybkie testy diagnostyczne** (np. CRP, strep-testy, testy combo antygenowe w kierunku grypy/COVID-19/RSV).<br><br>a. Znam ten typ strategii.<br>b. Regularnie stosuję tego typu strategie.<br>c. Czuję się pewnie w stosowaniu tego typu strategii.<br>d. Uważam, że ten typ strategii jest efektywny w ochronie antybiotyków. | **Do you agree with the following statements regarding these types of strategies?**<br><br>**Rapid diagnostic tests** (e.g., CRP, Strep tests, combo antigen tests for influenza/COVID-19/RSV).<br><br>a. I am familiar with this type of strategy.<br>b. I regularly use this type of strategy.<br>c. I feel confident using this type of strategy.<br>d. I believe this type of strategy is effective in antibiotic stewardship. |
| **Q15** | **Czy zgadza się Pan/Pani z zaprezentowanymi stwierdzeniami w odniesieniu do poniższych typów strategii?**<br><br>**Konkretne strategie komunikacji z pacjentami** o antybiotykach (np. odniesienie się bezpośrednio do oczekiwań, rozmowa o skutkach ubocznych, typowej długości trwania symptomów). | **Do you agree with the following statements regarding these types of strategies?**<br><br>**Specific communication strategies with patients regarding antibiotics** (e.g., addressing patient expectations, discussing side effects, typical symptom duration). |

| | | |
|---|---|---|
| | a. Znam ten typ strategii.<br>b. Regularnie stosuję tego typu strategie.<br>c. Czuję się pewnie w stosowaniu tego typu strategii.<br>d. Uważam, że ten typ strategii jest efektywny w ochronie antybiotyków. | a. I am familiar with this type of strategy.<br>b. I regularly use this type of strategy.<br>c. I feel confident using this type of strategy.<br>d. I believe this type of strategy is effective in antibiotic stewardship. |
| **Q16** | **Czy zgadza się Pan/Pani z zaprezentowanymi stwierdzeniami w odniesieniu do poniższych typów strategii?**<br><br>**Odroczone recepty na antybiotyki** (z datą „realizacji od”, w celu odroczenia rozpoczęcia antybiotykoterapii, np. podczas wyczekiwania na rozwój infekcji lub wyniki badań).<br><br>a. Znam ten typ strategii.<br>b. Regularnie stosuję tego typu strategie.<br>c. Czuję się pewnie w stosowaniu tego typu strategii.<br>d. Uważam, że ten typ strategii jest efektywny w ochronie antybiotyków. | **Do you agree with the following statements regarding these types of strategies?**<br><br>**Delayed antibiotic prescriptions** (with a "fill from" date to delay starting antibiotic therapy, e.g., while waiting to see how the infection develops or for test results).<br><br>a. I am familiar with this type of strategy.<br>b. I regularly use this type of strategy.<br>c. I feel confident using this type of strategy.<br>d. I believe this type of strategy is effective in antibiotic stewardship. |
| **Q17** | **W ostatnim sezonie infekcyjnym, jak często stosował/a Pan/Pani poniższe strategie u pacjentów z objawami infekcji układu oddechowego?** (Pytamy szacunkowo)<br><br>*(Opcje odpowiedzi: nigdy, rzadko (<50% pacjentów z objawami infekcji układu oddechowego), czasami (50-80% pacjentów z objawami infekcji układu oddechowego), często (>80% pacjentów z objawami infekcji układu oddechowego))*<br><br>a. Skale/kryteria kliniczne<br>b. Szybkie testy (np. strep-testy, testy CRP lub Combo)<br>c. Konkretne strategie komunikacji o antybiotykach<br>d. Odroczone recepty na antybiotyki | **In the last infection season, how often did you use the following strategies in patients with symptoms of respiratory tract infections?** (Please estimate)<br><br>*(Response options: Never / Rarely [<50% of patients with respiratory infection symptoms] / Sometimes [50–80% of patients with respiratory infection symptoms] / Often [>80% of patients with respiratory infection symptoms])*<br><br>a. Clinical scales/criteria<br>b. Rapid diagnostic tests (e.g., Strep tests, CRP, or Combo tests)<br>c. Specific communication strategies regarding antibiotics<br>d. Delayed antibiotic prescriptions |
| **Q18** | **Miejsce na komentarze związane z tymi strategiami** (opcjonalnie)<br>*(Pytanie otwarte)* | **Space for comments regarding these strategies** (optional)<br>*(Open-ended question)* |

#### Section 5. Views on implementation of AMS strategies

W tej sekcji pytamy o Pana/Pani poglądy związane z **wdrażaniem** działań w celu ochrony i optymalizacji stosowania antybiotyków i przeciwdziałania antybiotykooporności.

In this section, we ask about your views on implementing actions to promote antibiotic stewardship, optimise antibiotic use, and combat antimicrobial resistance.

| | | |
|---|---|---|
| **Q19** | **Który typ z obecnie stosowanych strategii ochrony antybiotyków powinnien być bardziej promowany i częściej stosowany w podstawowej opiece zdrowotnej w Polsce?**<br>[Pytamy tylko o strategie kierowane do lub stosowane przez lekarzy.]<br><br>*(Pytanie wielokrotnego wyboru)*<br><br>a. Krajowe programy i strategie<br>b. Wytyczne kliniczne<br>c. Kampanie informacyjne skierowane do lekarzy<br>d. Kursy, szkolenia, materiały edukacyjne dla lekarzy<br>e. Kryteria, skale, algorytmy kliniczne/diagnostyczne<br>f. Szybkie testy diagnostyczne<br>g. Odroczone recepty na antybiotyki<br>h. Szkolenia z efektywnej komunikacji z pacjentem o antybiotykach lub braku wskazań do stosowania antybiotykoterapii<br>i. Inne - Jakie? | **Which of the currently used antibiotic stewardship strategies should be more promoted and used more frequently in primary care in Poland?**<br>[We are asking only about strategies aimed at or used by physicians.]<br><br>*(Multiple-choice question)*<br><br>a. National programs and strategies<br>b. Clinical guidelines<br>c. Information campaigns targeted at physicians<br>d. Courses, training, and educational materials for physicians<br>e. Clinical/diagnostic criteria, scales, and algorithms<br>f. Rapid diagnostic tests<br>g. Delayed antibiotic prescriptions<br>h. Training in effective patient communication regarding antibiotics or the lack of indication for antibiotic therapy<br>i. Other – Please specify |
| **Q20** | **Który typ strategii mógłby pozytywnie wpłynąć na optymalizację przepisywania antybiotyków w podstawowej opiece zdrowotnej w Polsce?**<br>[Strategie te są stosowane i mają dowody efektywności w innych krajach.]<br><br>*(Pytanie wielokrotnego wyboru)*<br><br>a. Konkretny plan działania dla przychodni/praktyk lekarza rodzinnego związanych z optymalizacją antybiotyków<br>b. Liderzy opinii promujący optymalizację antybiotyków w medycynie rodzinnej | **Which type of strategy could positively impact the optimization of antibiotic prescribing in primary care in Poland?**<br>[These strategies are currently used and supported by evidence of effectiveness in other countries.]<br><br>*(Multiple-choice question)*<br><br>a. A specific action plan for clinics/family medicine practices related to antibiotic optimization<br>b. Opinion leaders promoting antibiotic optimization in family medicine |

| | | |
|---|---|---|
| | c. Monitorowanie i informacja zwrotna związana z liczbą przepisywanych antybiotyków przez daną placówkę, z porównaniem do innych podobnych placówek<br>d. Monitorowanie i informacja zwrotna związana z ilością przepisywanych antybiotyków przez indywidualnych lekarzy, z porównaniem do innych lekarzy lub średniej<br>e. Zachęty finansowe dla przychodni/praktyk lekarza rodzinnego związane z osiągnięciem ustalonych celów optymalizacji przepisywanych antybiotyków<br>f. Ulotki i materiały informacyjne, do których lekarz mógłby się odnieść w komunikacji z pacjentami<br>g. Inne - Jakie? | c. Audit and feedback on the volume of antibiotics prescribed by a given facility, compared to other similar facilities<br>d. Audit and feedback on the volume of antibiotics prescribed by individual physicians, compared to other physicians or the average<br>e. Financial incentives for clinics/family medicine practices tied to achieving established antibiotic optimization goals<br>f. Leaflets and educational materials that physicians can refer to when communicating with patients<br>g. Other – Please specify |
| **Q21** | **W Pana/Pani doświadczeniu, co najbardziej utrudnia wdrożenie strategii ochrony antybiotyków w podstawowej opiece zdrowotnej w Polsce?**<br><br>*(Pytanie wielokrotnego wyboru)*<br><br>a. Niedostateczna świadomość lekarzy jakie strategie wdrożyć<br>b. Brak priorytetu lub istotności ochrony antybiotyków<br>c. Brak odpowiedniego wsparcia we wdrażaniu strategii<br>d. Obciążenie ilością pracy i brak czasu w codziennej praktyce<br>e. Ograniczenia personalne<br>f. Ograniczenia sprzętowe<br>g. Inne - Jakie? | **In your experience, what are the main barriers to implementing antibiotic stewardship strategies in primary care in Poland?**<br><br>*(Multiple-choice question)*<br><br>a. Insufficient awareness among physicians regarding which strategies to implement<br>b. Lack of prioritization or perceived importance of antibiotic stewardship<br>c. Lack of adequate support in strategy implementation<br>d. Heavy workload and lack of time in daily clinical practice<br>e. Staffing/workforce constraints<br>f. Equipment limitations<br>g. Other – Please specify |
| **Q22** | **Co najbardziej pomaga lub pomogłyby wdrożyć strategie ochrony antybiotyków w podstawowej opiece zdrowotnej w Polsce?**<br><br>*(Pytanie wielokrotnego wyboru)*<br><br>a. Większa świadomość związana z potrzebą ochrony antybiotyków<br>b. Większa świadomość jakie strategie/działania wdrażać | **What most helps or would help to implement antibiotic stewardship strategies in primary care in Poland?**<br><br>*(Multiple-choice question)*<br><br>a. Greater awareness of the need for antibiotic stewardship<br>b. Greater awareness of which strategies/actions to implement |

| | Polski | English |
|---|---|---|
| | c. Nacisk na optymalizację stosowania antybiotyków jako priorytet dobrej opieki<br>d. Informacje i materiały dla przychodni i lekarzy wspierające wdrażanie odpowiednich strategii<br>e. Zasoby finansowe na wdrażanie strategii<br>f. Inne - Jakie? | c. Emphasis on optimizing antibiotic use as a priority of good care<br>d. Information and materials for clinics and physicians supporting the implementation of relevant strategies<br>e. Financial resources for strategy implementation<br>f. Other – Please specify |

#### Summary

| | | |
|---|---|---|
| **Q23** | **Miejsce na komentarze związane z tematem ankiety** (opcjonalnie)<br>*(Pytanie otwarte)* | **Space for comments related to the survey topic** (optional)<br>*(Open-ended question)* |

### Supplement 3: Flow diagram of the number of respondents for each survey section

#### Figure S1. Flow diagram of respondents through the survey

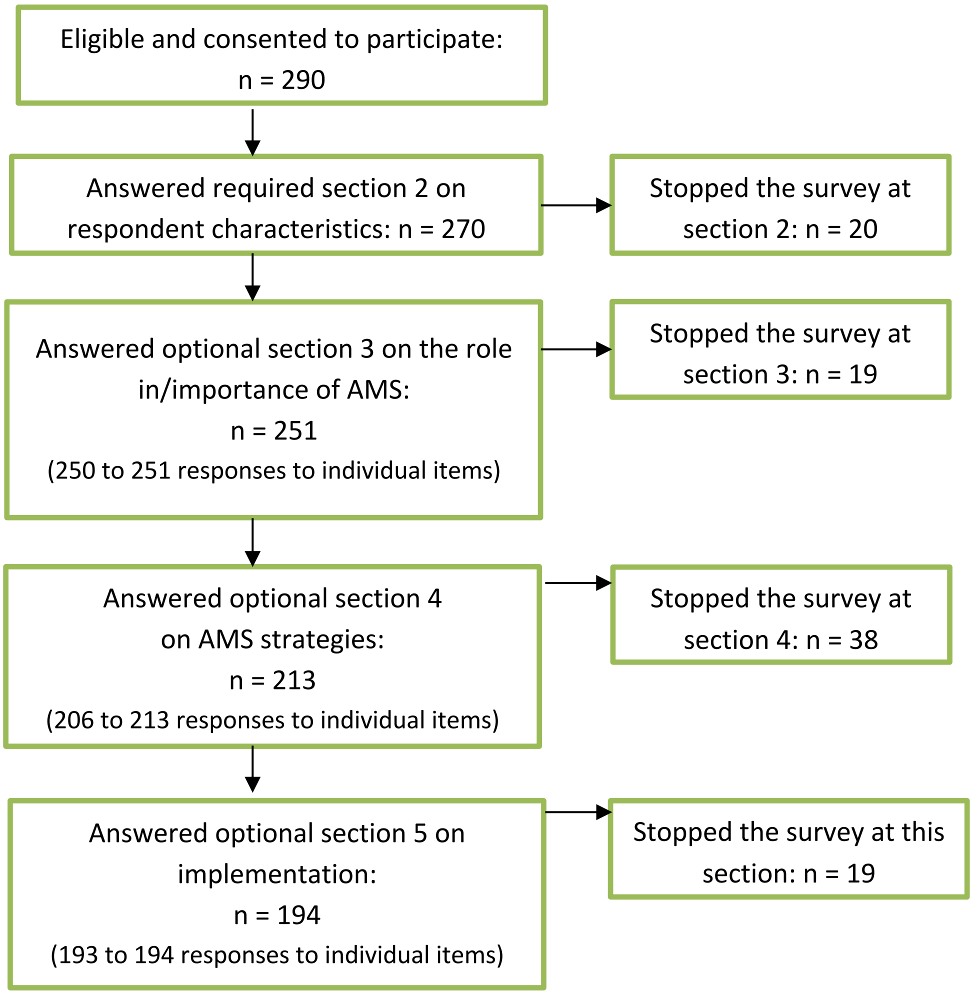

### Supplement 4: Additional respondent characteristics

#### Table S2. Region of Poland where the respondents typically work

| **Voivodeship (Province)** | **Number of respondents** (N=270) | **Proportion of respondents** (%) |
|---|---|---|
| Lower Silesian (*dolnośląskie*) | 36 | 13.3 |
| Kuyavian-Pomeranian (*kujawsko-pomorskie*) | 13 | 4.8 |
| Lublin (*lubelskie*) | 16 | 5.9 |
| Lubusz (*lubuskie*) | 4 | 1.5 |
| Łódź (*łódzkie*) | 22 | 8.1 |
| Lesser Poland (*małopolskie*) | 32 | 11.9 |
| Masovian (*mazowieckie*) | 31 | 11.5 |
| Opole (*opolskie*) | 7 | 2.6 |
| Subcarpathian (*podkarpackie*) | 14 | 5.2 |
| Podlaskie (*podlaskie*) | 10 | 3.7 |
| Pomeranian (*pomorskie*) | 20 | 7.4 |
| Silesian (*śląskie*) | 29 | 10.7 |
| Warmian-Masurian (*warmińsko-mazurskie*) | 8 | 3.0 |
| Greater Poland (*wielkopolskie*) | 24 | 8.9 |
| West Pomeranian (*zachodniopomorskie*) | 4 | 1.5 |
| Holy Cross (*świętokrzyskie*) | 0 | 0.0 |

**Characteristics of physicians in Poland:**

The National Chamber of Physicians in Poland (Naczelna Izba Lekarska) published the following statistical data about physicians in Poland (updated on 30.06.2026):

- Specialisation (dataset #04) – Number of professionally active specialists in Family Medicine: n = 12 441
- Gender and age groups (dataset #03) – Among professionally active physicians (any speciality or no speciality): men n = 66 222; women n = 98 234

The latest report (from 2022) on medical professionals from the Polish Office for Statistics (GUS) ("Zasoby kadrowe w wybranych zawodach medycznych na podstawie źródeł administacyjnych w latach 2019-2020") also reported the sex, age groups, and specialisations of all physicians, but did not provide characteristics of family physicians specifically.

We did not identify data reporting the characteristics of physicians working specifically in primary care.

### Supplement 5: Data tables

Q8. As a primary care physician, to what extent do you agree or disagree with the following statements?

| Items | Responses | n | % |
|---|---|---|---|
| I play an important role in antibiotic stewardship to prevent the development of drug resistance (n=251) | Strongly disagree | 12 | 4.78 |
| | Disagree | 7 | 2.79 |
| | Neither agree nor disagree | 8 | 3.19 |
| | Agree | 79 | 31.47 |
| | Strongly agree | 145 | 57.77 |
| Antibiotic stewardship and the optimal use of antibiotics are an important element of patient care in primary care (n=250) | Strongly disagree | 13 | 5.20 |
| | Disagree | 0 | 0.00 |
| | Neither agree nor disagree | 3 | 1.20 |
| | Agree | 48 | 19.20 |
| | Strongly agree | 186 | 74.40 |
| Antimicrobial resistance is a significant threat to patients in primary care (n=251) | Strongly disagree | 9 | 3.59 |
| | Disagree | 7 | 2.79 |
| | Neither agree nor disagree | 15 | 5.98 |
| | Agree | 77 | 30.68 |
| | Strongly agree | 143 | 56.97 |

Q9. Actions addressing AMR within national programmes and plans

| Items | Responses | n | % |
|---|---|---|---|
| I am familiar with this type of strategy (n=213) | Agree | 130 | 61.03 |
| | Partially agree | 47 | 22.07 |
| | Do not agree | 34 | 15.96 |
| | Not relevant | 2 | 0.94 |
| I regularly refer to this type of strategy (n=212) | Agree | 118 | 55.66 |
| | Partially agree | 54 | 25.47 |
| | Do not agree | 27 | 12.74 |
| | Not relevant | 13 | 6.13 |
| I believe this type of strategy is effective in AMS (n=212) | Agree | 91 | 42.92 |
| | Partially agree | 79 | 37.26 |
| | Do not agree | 21 | 9.91 |
| | Not relevant | 21 | 9.91 |

Q10. Clinical guidelines and recommendations related to the treatment of infections and antibiotic therapy in family medicine

| Items/questions | Responses | n | % |
|---|---|---|---|
| I am familiar with this type of strategy (n=213) | Agree | 206 | 96.71 |
| | Partially agree | 5 | 2.35 |
| | Do not agree | 2 | 0.94 |
| | Not relevant | 0 | 0.00 |

| | | | |
|---|---|---|---|
| I regularly refer to this type of strategy (n=213) | Agree | 190 | 89.20 |
| | Partially agree | 16 | 7.51 |
| | Do not agree | 6 | 2.82 |
| | Not relevant | 1 | 0.47 |
| I believe this type of strategy is effective in AMS (n=212) | Agree | 152 | 71.70 |
| | Partially agree | 50 | 23.58 |
| | Do not agree | 9 | 4.25 |
| | Not relevant | 1 | 0.47 |

Q11. Information and educational campaigns related to antibiotics, AMR, or the treatment of infections

| Items/questions | Responses | n | % |
|---|---|---|---|
| I am familiar with this type of strategy (n=213) | Agree | 87 | 40.85 |
| | Partially agree | 50 | 23.47 |
| | Do not agree | 74 | 34.74 |
| | Not relevant | 2 | 0.94 |
| I believe this type of strategy is effective in AMS (n=212) | Agree | 55 | 25.94 |
| | Partially agree | 67 | 31.60 |
| | Do not agree | 45 | 21.23 |
| | Not relevant | 45 | 21.23 |

Q12. Education and training related to antibiotic therapy, drug resistance, and infectious diseases

| Items/questions | Responses | n | % |
|---|---|---|---|
| I regularly participate in such training (n=213) | Agree | 116 | 54.46 |
| | Partially agree | 70 | 32.86 |
| | Do not agree | 24 | 11.27 |
| | Not relevant | 3 | 1.41 |
| Such training has increased my knowledge and skills (n=213) | Agree | 144 | 67.61 |
| | Partially agree | 43 | 20.19 |
| | Do not agree | 6 | 2.82 |
| | Not relevant | 20 | 9.39 |
| I believe this type of strategy is effective in AMS (n=212) | Agree | 138 | 65.09 |
| | Partially agree | 56 | 26.42 |
| | Do not agree | 7 | 3.30 |
| | Not relevant | 11 | 5.19 |

Q12a. In the past year, how often did you participate in the following forms of continuing education related to AMS or infectious diseases?

| Items | Responses | n | % |
|---|---|---|---|
| Training courses or webinars by the Polish Society of Family Medicine (PTMR) (n=213) | Never | 39 | 18.31 |
| | Rarely (1-2 times) | 98 | 46.01 |

| | | | |
|---|---|---|---|
| | Often (3-4 times) | 64 | 30.05 |
| | Very often (5+ times) | 12 | 5.63 |
| Training courses or specialised sessions at conferences (n=211) | Never | 39 | 18.48 |
| | Rarely (1-2 times) | 100 | 47.39 |
| | Often (3-4 times) | 63 | 29.86 |
| | Very often (5+ times) | 9 | 4.27 |
| Other training courses (n=207) | Never | 69 | 33.33 |
| | Rarely (1-2 times) | 83 | 40.10 |
| | Often (3-4 times) | 43 | 20.77 |
| | Very often (5+ times) | 12 | 5.80 |

Q12b. In the past year, how often did you use the following forms of continuing education related to AMS or infectious diseases?

| **Items** | **Responses** | **n** | **%** |
|---|---|---|---|
| Educational materials by the Polish Society of Family Medicine (PTMR) (n=212) | Never | 35 | 16.51 |
| | Rarely (1-2 times) | 90 | 42.45 |
| | Often (3-4 times) | 68 | 32.08 |
| | Very often (5+ times) | 19 | 8.96 |
| Reading specialised articles (medical, scientific) (n=212) | Never | 17 | 8.02 |
| | Rarely (1-2 times) | 46 | 21.70 |
| | Often (3-4 times) | 94 | 44.34 |
| | Very often (5+ times) | 55 | 25.94 |
| Other forms of continuing education (n=207) | Never | 78 | 37.68 |
| | Rarely (1-2 times) | 48 | 23.19 |
| | Often (3-4 times) | 52 | 25.12 |
| | Very often (5+ times) | 29 | 14.01 |

Q13. Clinical scales/criteria and diagnostic algorithms related to infection diagnosis and antibiotic therapy

| **Items** | **Responses** | **n** | **%** |
|---|---|---|---|
| I am familiar with this type of strategy (n=213) | Agree | 204 | 95.77 |
| | Partially agree | 5 | 2.35 |
| | Do not agree | 4 | 1.88 |
| | Not relevant | 0 | 0.00 |
| I regularly use this type of strategy (n=211) | Agree | 177 | 83.89 |
| | Partially agree | 28 | 13.27 |
| | Do not agree | 6 | 2.84 |
| | Not relevant | 0 | 0.00 |
| I feel confident using this type of strategy (n=211) | Agree | 159 | 75.36 |
| | Partially agree | 40 | 18.96 |
| | Do not agree | 11 | 5.21 |
| | Not relevant | 1 | 0.47 |
| I believe this type of strategy is effective in AMS (n=212) | Agree | 139 | 65.57 |

| | | | |
|---|---|---|---|
| | Partially agree | 63 | 29.72 |
| | Do not agree | 9 | 4.25 |
| | Not relevant | 1 | 0.47 |

Q14. Rapid diagnostic tests

| Items | Responses | n | % |
|---|---|---|---|
| I am familiar with this type of strategy (n=212) | Agree | 208 | 98.11 |
| | Partially agree | 3 | 1.42 |
| | Do not agree | 1 | 0.47 |
| | Not relevant | 0 | 0.00 |
| I regularly use this type of strategy (n=213) | Agree | 200 | 93.90 |
| | Partially agree | 5 | 2.35 |
| | Do not agree | 7 | 3.29 |
| | Not relevant | 1 | 0.47 |
| I feel confident using this type of strategy (n=213) | Agree | 200 | 93.90 |
| | Partially agree | 8 | 3.76 |
| | Do not agree | 4 | 1.88 |
| | Not relevant | 1 | 0.47 |
| I believe this type of strategy is effective in AMS (n=211) | Agree | 172 | 81.52 |
| | Partially agree | 35 | 16.59 |
| | Do not agree | 4 | 1.90 |
| | Not relevant | 0 | 0.00 |

Q15. Specific communication strategies with patients regarding antibiotics

| Items | Responses | n | % |
|---|---|---|---|
| I am familiar with this type of strategy (n=213) | Agree | 195 | 91.55 |
| | Partially agree | 9 | 4.23 |
| | Do not agree | 8 | 3.76 |
| | Not relevant | 1 | 0.47 |
| I regularly use this type of strategy (n=212) | Agree | 184 | 86.79 |
| | Partially agree | 18 | 8.49 |
| | Do not agree | 9 | 4.25 |
| | Not relevant | 1 | 0.47 |
| I feel confident using this type of strategy (n=213) | Agree | 152 | 71.36 |
| | Partially agree | 48 | 22.54 |
| | Do not agree | 12 | 5.63 |
| | Not relevant | 1 | 0.47 |
| I believe this type of strategy is effective in AMS (n=212) | Agree | 101 | 47.64 |
| | Partially agree | 95 | 44.81 |
| | Do not agree | 13 | 6.13 |
| | Not relevant | 3 | 1.42 |

Q16. Delayed antibiotic prescriptions

| Items | Responses | n | % |
|---|---|---|---|
| I am familiar with this type of strategy (n=213) | Agree | 135 | 63.98 |
| | Partially agree | 23 | 10.90 |
| | Do not agree | 51 | 24.17 |
| | Not relevant | 2 | 0.95 |
| I regularly use this type of strategy (n=212) | Agree | 32 | 15.31 |
| | Partially agree | 57 | 27.27 |
| | Do not agree | 112 | 53.59 |
| | Not relevant | 8 | 3.83 |
| I feel confident using this type of strategy (n=213) | Agree | 53 | 25.73 |
| | Partially agree | 45 | 21.84 |
| | Do not agree | 70 | 33.98 |
| | Not relevant | 38 | 18.45 |
| I believe this type of strategy is effective in AMS (n=212) | Agree | 42 | 20.00 |
| | Partially agree | 63 | 30.00 |
| | Do not agree | 90 | 42.86 |
| | Not relevant | 15 | 7.14 |

Q17. In the last infection season, how often did you use the following strategies in patients with symptoms of RTIs?

| Items | Responses | n | % |
|---|---|---|---|
| Clinical scales/criteria (n=213) | Never | 4 | 1.88 |
| | Rarely [<50% of patients with respiratory infection symptoms] | 24 | 11.27 |
| | Sometimes [50-80% of patients with respiratory infection symptoms] | 44 | 20.66 |
| | Often [>80% of patients with respiratory infection symptoms] | 141 | 66.20 |
| Rapid diagnostic tests (e.g., Strep tests, CRP, or Combo tests) (n=213) | Never | 4 | 1.88 |
| | Rarely [<50% of patients with respiratory infection symptoms] | 9 | 4.23 |
| | Sometimes [50-80% of patients with respiratory infection symptoms] | 45 | 21.13 |
| | Often [>80% of patients with respiratory infection symptoms] | 155 | 72.77 |
| Specific communication strategies regarding antibiotics (n=213) | Never | 6 | 2.82 |
| | Rarely [<50% of patients with respiratory infection symptoms] | 23 | 10.80 |

| | | | |
|---|---|---|---|
| | Sometimes [50-80% of patients with respiratory infection symptoms] | 76 | 35.68 |
| | Often [>80% of patients with respiratory infection symptoms] | 108 | 50.70 |
| Delayed antibiotic prescriptions(n=213) | Never | 110 | 51.64 |
| | Rarely [<50% of patients with respiratory infection symptoms] | 78 | 36.62 |
| | Sometimes [50-80% of patients with respiratory infection symptoms] | 17 | 7.98 |
| | Often [>80% of patients with respiratory infection symptoms] | 8 | 3.76 |

Q19. Which of the currently used AMS strategies should be more promoted and used in primary care in Poland?

| **Items** | **N (n=194)** | **% (100%)** |
|---|---|---|
| Rapid diagnostic tests | 144 | 74.2% |
| Clinical guidelines | 142 | 73.2% |
| Clinical criteria, scales, algorithms | 129 | 66.5% |
| Training and educational resources for clinicians | 127 | 65.5% |
| Training in communication skills about antibiotics | 96 | 49.5% |
| Informational campaigns directed at clinicians | 87 | 44.8% |
| National programmes and strategies | 65 | 33.5% |
| Delayed antibiotic prescriptions | 14 | 7.2% |
| Other | 13 | 6.7% |

Q20. Which of the potential AMS strategies could positively impact on antibiotic prescribing in primary care in Poland?

| **Items** | **N (n=194)** | **% (100%)** |
|---|---|---|
| Specific action plan for general practices related to optimising antibiotic prescribing | 138 | 71.1% |
| Leaflets and informational resources that a clinician could refer to when communicating with patients | 103 | 53.1% |
| Monitoring and feedback on the number of prescribed antibiotics by individual physicians, with a comparison to peers or to an average | 98 | 50.5% |
| Monitoring and feedback on the number of prescribed antibiotics by general practices, with a comparison to other practices | 94 | 48.5% |
| Financial incentives for general practices linked to achieving pre-set goals for optimising antibiotic prescribing | 67 | 34.5% |
| Opinion leaders promoting the optimisation of antibiotic prescribing in family medicine | 36 | 18.6% |
| Other | 6 | 3.1% |

Q21. In your experience, what are the main barriers to implementing antibiotic stewardship strategies in primary care in Poland?

| Items | N (n=193) | % (100%) |
|---|---|---|
| Heavy workload and lack of time in daily clinical practice | 121 | 62.7 |
| Insufficient awareness among physicians regarding which strategies to implement | 121 | 62.7 |
| Lack of prioritization or perceived importance of AMS | 89 | 46.1 |
| Staffing constraints | 66 | 34.2 |
| Lack of adequate support in strategy implementation | 51 | 26.4 |
| Other | 38 | 19.7 |
| Equipment limitations | 25 | 13.0 |

Q22. What most helps or would help to implement antibiotic stewardship strategies in primary care in Poland?

| Items | N (n=194) | % (100%) |
|---|---|---|
| Greater awareness of the need for AMS | 128 | 66.0 |
| Emphasis on optimizing antibiotic use as a priority of good care | 113 | 58.2 |
| Greater awareness of which strategies/actions to implement | 95 | 49.0 |
| Information and materials for clinics and physicians supporting the implementation of relevant strategies | 95 | 49.0 |
| Financial resources for strategy implementation | 60 | 30.9 |
| Other | 9 | 4.6 |
